# Quantifying factors that affect polygenic risk score performance across diverse ancestries and age groups for body mass index

**DOI:** 10.1101/2022.05.27.22275647

**Authors:** Daniel Hui, Brenda Xiao, Ozan Dikilitas, Robert R. Freimuth, Marguerite R. Irvin, Gail P. Jarvik, Leah Kottyan, Iftikhar Kullo, Nita A. Limdi, Cong Liu, Yuan Luo, Bahram Namjou, Megan J. Puckelwartz, Daniel Schaid, Hemant Tiwari, Wei-Qi Wei, Shefali Verma, Dokyoon Kim, Marylyn D. Ritchie

## Abstract

Polygenic risk scores (PRS) have led to enthusiasm for precision medicine. However, it is well documented that PRS do not generalize across groups differing in ancestry or sample characteristics e.g., age. Quantifying performance of PRS across different groups of study participants, using genome-wide association study (GWAS) summary statistics from multiple ancestry groups and sample sizes, and using different linkage disequilibrium (LD) reference panels may clarify factors limiting PRS transferability. To evaluate these factors in the PRS generation process, we generated body mass index (BMI) PRS (PRS_BMI_) in the Electronic Medical Records and Genomics network (N=75,661). Analyses were conducted in two ancestry groups (European and African) and three age ranges (adult, teenagers, and children). For PRS_BMI_ calculations, we evaluated five LD reference panels and three GWAS summary statistics of varying sample size and ancestry. PRS_BMI_ performance increased for both African and European ancestry individuals using cross-ancestry GWAS summary statistics compared to European-only summary statistics (6.3% and 3.7% relative R^2^ increase, respectively, p_African_=0.038, p_European_=6.26×10^−4^). The effects of LD reference panels were more pronounced in African ancestry study datasets. PRS_BMI_ performance degraded in children; R^2^ was less than half of teenagers or adults. The effect of GWAS summary statistics sample size was small when modeled with the other factors. We also explored clinical comorbidities associated with the PRS_BMI_ and identified associations with type 2 diabetes and coronary atherosclerosis. This study quantifies effects that ancestry, GWAS summary statistic sample size, and LD reference panel have on PRS performance, especially in cross-ancestry and age-specific analyses.

## Introduction

Polygenic risk scores (PRS) provide individualized genetic estimates of a phenotype by aggregating genetic effects across hundreds or thousands of loci, typically from genome-wide association studies (GWAS). PRS are potentially a powerful source of increased prediction performance, even when combined with familial history (1,2). However, in recent years it has become increasingly apparent that performance of PRS is substantially reduced when the ancestry of the individuals in whom prediction is being done differs from the ancestry of the individuals from the GWAS used to generate SNP weights used for PRS construction. For instance, when using GWAS from European ancestry individuals, the prediction accuracy of polygenic scores in individuals of African or Hispanic/Latino ancestry have a relative performance of 25% and 65% compared to performance in European ancestry individuals (3). Additionally, evidence exists suggesting that for some traits, such as adiposity traits, this disparity may be further exacerbated by environmental, demographic, or social risk factors (including age, physical activity, smoking status, and alcohol use (4–7)). For example, differences in the genetic architecture of body mass index (BMI) has been shown to differ between age groups (8–11). Thus, the performance of PRS for BMI is also affected by the age of the individuals used in the GWAS and the test dataset (12). Broad-sense heritability estimates for BMI in adults range from 40%-90% when estimated in adults of different cohorts even of homogeneous ancestry (13) – even if heritability estimates are similar across populations, genetic architecture and enrichment for variants in different functional categories may still differ (14,15).

Several outstanding questions surrounding PRS, especially within the context of adiposity traits and BMI, warrant further investigation. For instance, when cross-ancestry summary statistics (i.e., those including individuals of multiple ancestry groups) are available, can they be used to improve prediction performance in individuals from one or more different ancestry groups. We need a more thorough evaluation of the potential prediction performance gain (or loss) in African ancestry individuals when cross-ancestry GWAS summary statistics are used to estimate the SNP weights. In addition, we need to improve our understanding of the impact of the composition of the linkage disequilibrium (LD) reference panel in combination with cross-ancestry GWAS summary statistics on the PRS prediction performance. For prediction of BMI specifically, how does prediction performance differ for individuals in different age groups, especially those who are not adults (i.e., less than age 18). Additionally, how much do these different variables impact the PRS performance when considered together; and how much do these differences affect associations with clinical comorbidities of obesity. Finally, the degree to which increased GWAS sample size increases prediction performance regardless of these other factors is important to determine.

We comprehensively investigated the influence of these factors on the performance of PRS using the Electronic Medical Records and Genomics (eMERGE) Network dataset. eMERGE is a prospective study that combines participants from multiple electronic health record (EHR) linked biobanks (16). In the present study, we included 75,661 individuals of diverse ancestry and age (14% African ancestry, 55% female, and 12% children age <13). These individuals were from the eMERGE III imputed array dataset (N=83,717) (dbGaP Study: phs001584.v2.p2), estimated European or African ancestry, and had BMI measurements available. For these analyses, we used published BMI GWAS summary statistics from the GIANT (Genetic Investigation of ANthropometric Traits) consortium, an international consortium that primarily studies anthropometric traits, which included participants (max N=339,224, mean N per variant=226,960) from European, African, and Asian ancestry groups (17). We also used summary statistics from a European ancestry BMI GWAS (18) in the UK Biobank (UKBB) individuals (N=339,721), which was conducted using both the full sample size of the European ancestry UKBB, as well as after down sampling to the same number of individuals in the GIANT GWAS. This comparison allowed us to better evaluate whether it was the ancestry composition or the sample size of the dataset where the GWAS summary statistics were derived that affected the results of the PRS performance. We calculated PRS for BMI (PRS_BMI_) across 90 different combinations of analyses (described more in Methods) – 6 different groupings based on ancestry and age, five different LD reference panels (of varying ancestry and from three different cohorts), and the three mentioned sets of summary statistics. We then statistically compared the different sets of analyses to see what factors most influence the PRS_BMI_ performance across various groupings of individuals based on ancestry and age. Lastly, we also tested the association of the best performing PRS_BMI_ with common comorbidities across ancestry groups to identify the clinical relevance of the PRS_BMI_ in phenotypes derived from an Electronic Health Record (EHR). Investigation of these variables elucidates our understanding of the factors that affect PRS performance and transferability across ancestries and populations, especially within the context of BMI.

## Methods

### Overall study design

The electronic Medical Records and Genomics (eMERGE) network dataset is a prospective study that combines participants from multiple electronic health record (EHR) linked biobanks. In this study, we included 75,661 individuals with available genetic and phenotypic data. The individuals in the eMERGE dataset include multiple ancestry groups and a large age distribution (14% genetically inferred African ancestry, 19% less than age 18, Figure 1). Briefly, we calculated PRS_BMI_ for all individuals, for each combination where the following elements of the model varied: 1) LD panels that differed in ancestry, 2) GWAS summary statistics with variable ancestry composition, and 3) GWAS summary statistics for two different sample sizes. The details for each of these are provided more below. We then split the data by ancestry and age group, and statistically compared PRS_BMI_ performance between all the different groups – in total, 90 sets of PRS_BMI_ were compared. We first estimated the effect and significance of each variable (i.e., ancestry of GWAS summary statistics and test data, LD panel ancestry, size of GWAS summary statistics, and age of test individuals) on PRS performance. Next, we estimated how much each variable affects PRS_BMI_ performance when all are modeled together, and finally we analyzed the potential clinical associations by testing the PRS_BMI_ for association with common comorbid conditions from the EHR. For the primary results related to LD panel or ancestry of summary statistics and test data, we restricted analyses to adults as the other age groups were limited in sample size.

**Figure 1.**
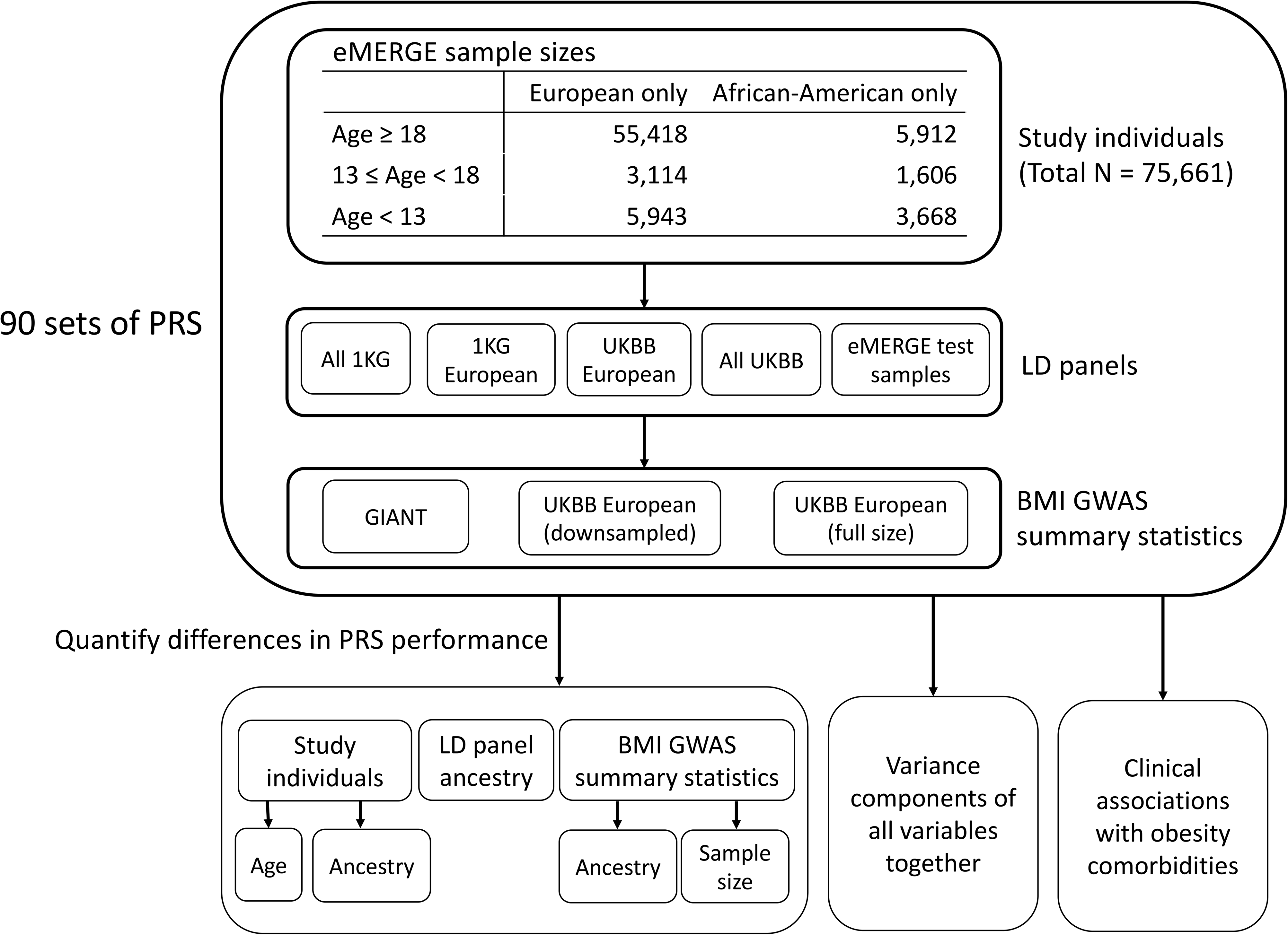
Flowchart of project. Max size of LD panel was 5,000 individuals. UK Biobank (UKBB) European GWAS summary statistics were downsampled to the mean sample size per variant of GIANT (N=226,960), full size of UKBB European was N=377,921. 1000 Genomes is abbreviated as 1KG.

### Summary statistics to generate PRS_BMI_

We obtained published GWAS summary statistics from the GIANT consortium (17) to use as one set of BMI GWAS summary statistics. Up to 322,154 adults of European ancestry, as well as an additional 17,072 adults of non-European ancestry (adults of African, East Asian, and South Asian ancestry), were included in the GIANT GWAS analysis.

For the second set of summary statistics, we performed a GWAS in the individuals of European ancestry from the UK Biobank (UKBB). Individuals were first filtered by low quality samples (sex mismatch between genetically inferred and self-reported, variant missingness >5%), relatedness (no 2nd degree relatives or higher), and within the White British ancestry subset (with these individuals being defined by UKBB and selected based on self-reports and genetically determined ancestry) (18) – 377,921 individuals initially remained. Variants were filtered on imputation quality score (using the INFO metric (19)) > 0.30, and minor allele frequency >1% within this subset of individuals. In addition, we generated a second set of GWAS summary statistics from the UKBB, where we randomly down-sampled individuals to the sample size in the GIANT GWAS dataset (N=226,960). In each UKBB GWAS, data processing and modeling were performed similarly as in the GIANT GWAS – summary statistics were calculated using linear regression, with age, age^2^, sex, and the first 5 genetic principal components (PCs) included as covariates. BMI, defined as weight in kilograms divided by squared height in meters, was first inverse-rank normal transformed.

After calculation of BMI GWAS summary statistics in each of the two datasets of UKBB individuals of European ancestry, we harmonized variants across all datasets used (UKBB, eMERGE, GIANT, and 1000 Genomes Phase 3). For the remainder of downstream analyses, we kept only those variants that were present in all datasets, and additionally excluded any strand-ambiguous SNPs (alleles A/T or C/G), and retained only biallelic variants; in total, 2,014,457 variants were retained for analyses.

### LD reference panels

Five different LD reference panels were used for each set of PRS_BMI_ calculations: 1) all of 1000 Genomes (1KG_All_) (N=2,504), 2) 1000 Genomes European ancestry (1KG_EUR_) (N=503), 3) 5,000 randomly selected European ancestry individuals from the UK Biobank (UKBB_EUR_), 4) 5,000 randomly selected individuals from all of UK Biobank (UKBB_All_), and 5) up to 5,000 randomly selected individuals from the test dataset for which PRS_BMI_ were being calculated for (referred to as test data henceforward).

### Statistical methods

#### PRS software

PRS_BMI_ were calculated using pruning and thresholding method via PRSice v2.1.9 (20). Default parameters were used in all analyses (clumping performed in 250 kb windows using an R^2^ of 0.1, p-value step size of 0.00005).

#### Statistical comparisons

Partial R^2^ for PRS_BMI_ was calculated by subtracting the R^2^ using a model with only the covariates from the R^2^ of the model using the covariates and the PRS_BMI_ (the default option in PRSice). P-values for the differences between model performance from all different iterations were calculated by using the Wilcoxon rank-sum test between the distributions of the squared residuals of the different models – for comparisons between the same set of individuals, the paired Wilcoxon rank-sum test was used. When testing which of the five LD panels performed the best, we used a Bonferroni-corrected threshold of 0.05/10 = 0.005 (ten comparisons between five LD panels). When comparing the best performing PRS_BMI_ across ancestries and summary statistics using their best LD panel, we used a Bonferroni threshold of 0.05/25 = 0.002 (25 comparisons between the five LD panels used).

#### Proportion of variance explained by each individual variable

We modeled all evaluated features together in the following linear regression model:

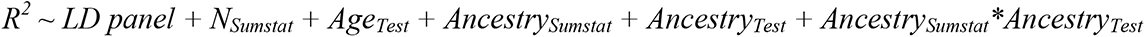

Where the *Sumstat* subscript is defined as a set of GWAS summary statistics, and the *Test* subscript is defined as a set of test individuals that PRS prediction is being assessed in. We quantified the variance in R^2^ that could be explained by each of these different variables using type II sum of squares from ANOVA. The sum of squares of variables involving ancestry were summed together; an interaction term between summary statistics ancestry and test data ancestry was included to identify whether the ancestry of summary statistics and test data matched.

#### Association of PRS_BMI_ with comorbidities

We selected the ten most frequent Phecodes (21) from the EHR data in the eMERGE dataset (which includes obesity as a positive control) to test their association with the PRS_BMI_ (S Table 2). For each Phecode, individuals were classified as a case for the condition if there was at least one occurrence of the respective Phecode in their EHR record; individuals were classified as a control for that condition if there was no occurrence of the Phecode. This classification is a rule-of-one instance of a Phecode to define case status. For each eMERGE ancestry subgroup, we selected the best performing PRS_BMI_ i.e., the PRS_BMI_ with the highest R^2^ and tested the association of the PRS_BMI_ with the most prevalent conditions using a logistic regression model. PRS_BMI_ was first mean-centered and standard deviation was set to 1. Sex, age, age^2^, and the first five genetic PCs were included as covariates.

### Data visualization

The ‘ggplot2’ R package was used for plotting, with the ‘geom_signif’ package used to include significance bars. The association results were plotted using PheWAS-View (22).

## Results

### Effect of LD panel

For adults of African ancestry, when using the down-sampled UKBB GWAS summary statistics, using either cross-ancestry or African ancestry LD panels significantly improved PRS_BMI_ performance compared to European ancestry LD panels (Figure 2). When using the UKBB summary statistics, the top PRS_BMI_ R^2^ was 0.0140 using the test data as LD panel, while the second-best performing LD panel (UKBB European) had an R^2^ of 0.0109 (p = 4.94×10^−20^). When using the GIANT summary statistics, the top PRS_BMI_ R^2^ was 0.0149 using 1KG_All_ as the reference panel. The PRS_BMI_ calculated using the best European ancestry panel (1KG_EUR_) resulted in a R^2^ of 0.141, but this difference between these two reference panels was not Bonferroni significant (p = 0.037). However, the 1KG_all_ LD panel performed significantly better than the two UKBB LD panels (UKBB_All_: R^2^ = 0.134, p = 3.65×10^−5^; UKBB European: R^2^ = 0.0128, p = 3.65×10^−9^). The test data LD panel performed the second-best with an R^2^ of 0.0142, and significantly outperformed the UKBB European LD panel (p = 4.78×10^−5^).

**Figure 2.**
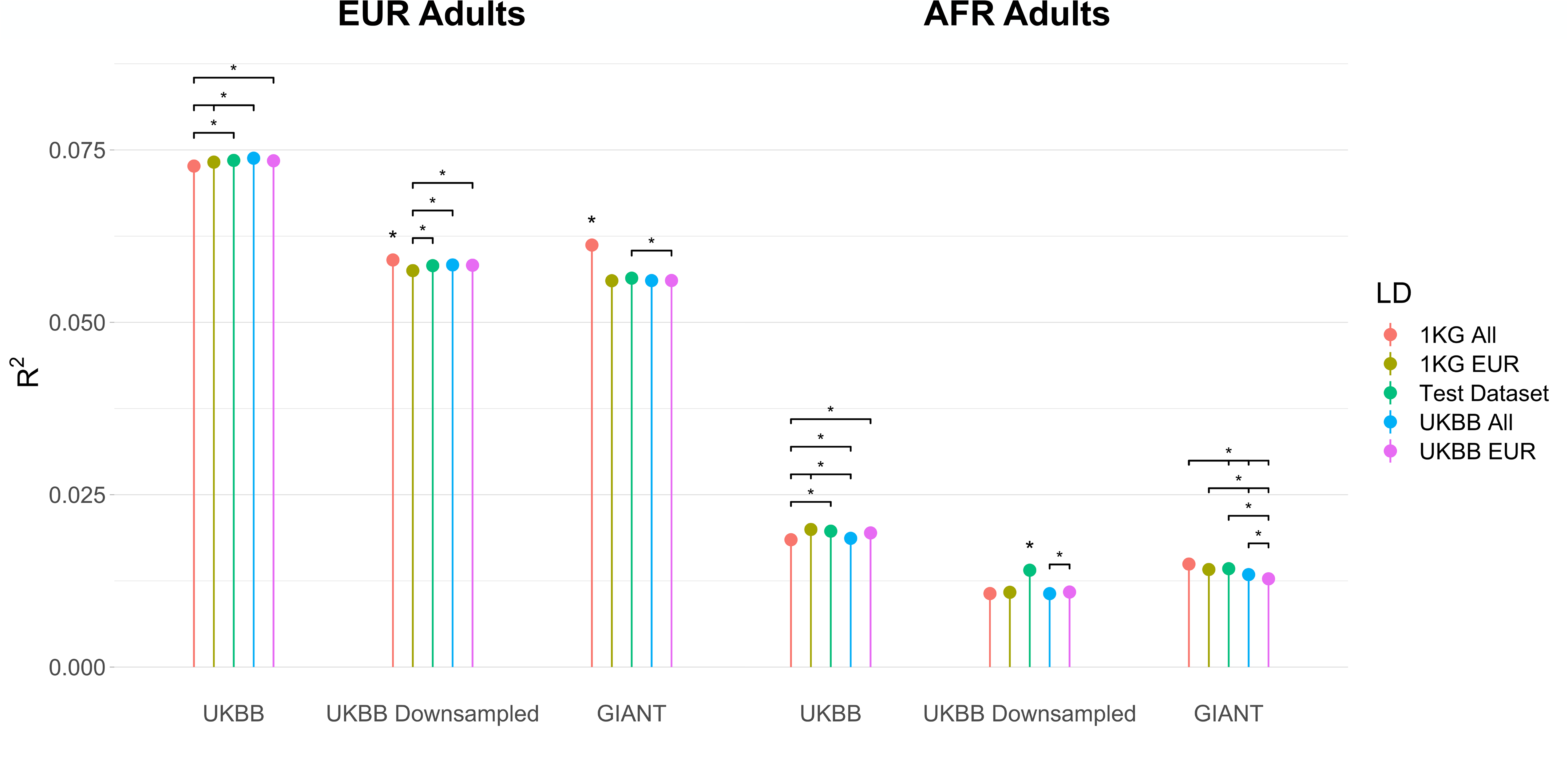
PRS R^2^ values across all runs in adults. Asterisks without bars indicate significantly different R^2^ values between the other 4 LD panels used. Bars are present for significant differences between specific R^2^ values.

For adults of European ancestry, we observed more significant differences in performance when using the GIANT summary statistics compared to the down-sampled UKBB summary statistics. The 1KG_All_ LD panel performed the best with a R^2^ of 0.0612. It also significantly outperformed all other LD panels (1KG_EUR_: R^2^ = 0.056, p = 5.54×10^−104^; Test data: R^2^ = 0.0564, p = 6.50×10^−67^; UKBB_All_: R^2^ = 0.0561, 8.09×10^−107^; UKBB_EUR_: R^2^ = 0.0561, p = 3.02×10^−77^). We note that this increase was larger when using the GIANT summary statistics but was still present when using the UKBB summary statistics. When using the UKBB summary statistics, the choice of LD panel had a much smaller impact on prediction performance. While the 1KG_All_ LD panel performed the best, the difference in performance was much less significant between the next best performing LD panel (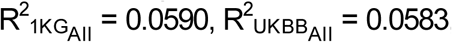, p = 3.48×10^−4^). The difference between the best and worst performing scores – LD panel using 1KG all versus 1KG European – was also much less significant (p= 1.15×10^−12^). These results suggest that the choice of LD panel particularly matters when calculating PRS_BMI_ using cross-ancestry GWAS, or for African ancestry individuals when the GWAS summary statistics are derived from European ancestry individuals.

However, we did observe a slight decrease in the impact of the choice of LD panel when using the full UKBB summary statistics for adults – again, the largest differences were observed in adults of African ancestry, but differences in performance across LD panels were not as significant. The test LD panel performed second best with the 1KG_EUR_ LD panel performing best (R^2^_Test_ = 0.0197, 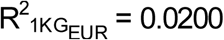, p = 0.18). The 1KG_All_ LD panel was the worst performing LD panel with an R^2^ of 0.185, and this difference between the 1KG_EUR_ LD panel was significant after multiple hypothesis correction (p = 5.08×10^−7^).

### Effect of summary statistics and ancestry of test data

As expected, the R^2^ values of the PRS_BMI_ were significantly higher when calculated for European ancestry adults than adults of African ancestry, even when using the cross-ancestry GIANT summary statistics (Figure 2). When using the GIANT summary statistics, the best performing PRS_BMI_ in adults of European ancestry had an R^2^ of 0.0612, which was significantly higher than the R^2^ from the best performing PRS_BMI_ in African ancestry adults (R^2^ = 0.0149, p < 4.9×10^−324^).

In African ancestry adults, the R^2^ when using the GIANT summary statistics was higher than the R^2^ when using the down-sampled UKBB summary statistics with their respective best LD panel (GIANT (1KG_All_ LD panel): R^2^ = 0.0149, UKBB (test data LD panel): R^2^ = 0.140; p = 0.038). This difference was not statistically significant after multiple hypothesis correction. However, the GIANT summary statistics with the 1KG_All_ LD panel did significantly outperform the UKBB summary statistics with all other LD panels. When keeping the LD panel constant, the PRS_BMI_ calculated using the GIANT summary statistics resulted in higher R^2^ than using the UKBB summary statistics for all LD panels except for the test data LD panel, and this difference was statistically significant for the 1KG_All_ (p = 1.55×10^−33^), 1KG_EUR_ (p = 6.78×10^−18^), and UKBB_All_ (p = 1.28×10^−15^) LD panels. Somewhat surprisingly, we observed higher R^2^ values for European ancestry adults when using the cross-ancestry GIANT summary statistics versus the down-sampled European UKBB summary statistics (R^2^_GIANT_ = 0.0612 versus R^2^_UKBB_ = 0.0590), with this difference being statistically significant (p = 6.26×10^−4^); the best performing LD panel for both set of summary statistics was 1KG_All_.

We also compared prediction performance in all individuals using the full (N=377,921) European UKBB GWAS versus the European UKBB GWAS down-sampled to GIANT’s sample size (N=226,960) (Figure 2, S Table 1). For consistency, UKBB European individuals were used for the European test ancestry comparisons, and for the African ancestry comparisons the test sets (i.e., African ancestry LD panels) were used as LD panels. Unanimously across test ancestry and age groups, we observed higher and statistically significant increases in R^2^.

### Prediction performance across different age groups

Across different ancestries and summary statistics, we broadly observed similar R^2^ values for adults and teenagers, with substantially reduced performance in children (S Figure 1). R^2^ values in children were consistently less than half of that in adults and teenagers, with differences in R^2^ values for adults and teenagers being minimal (except in the case of African ancestry individuals using the GIANT summary statistics, with teenagers having more than double the R^2^ of adults). Somewhat surprisingly, teenagers consistently had higher R^2^ than adults across all analyses, although these differences were much less significant than those compared with children.

### Proportion of variance explained by each assessed factor

While we observed significant differences due to ancestry, age, and number of individuals used to calculate summary statistics, we aimed to quantify the effect of these different variables on PRS_BMI_ performance when considered together (Table 1). We observed that 89.5% of the variance in PRS_BMI_ R^2^ could be explained using these variables, indicating that the majority of the effects of LD panel, ancestry, age, and sample size could be explained through linear relationships with PRS_BMI_ R^2^. In the context of these comparisons, the ancestry of the summary statistics or test data accounts for 55.1% of the variance explained in PRS_BMI_ R^2^. Choice of LD panel and age of test individuals accounted for similar amounts of variance explained in PRS_BMI_ R^2^ (16.5% and 15.9%, respectively), while the number of individuals used to calculate the GWAS summary statistics only accounted for 1.9% of variance explained in PRS_BMI_ R^2^. Per previous sections, while number of individuals used for summary statistics resulted in significant differences in PRS_BMI_ performance, its overall impact when modeled jointly with all the other factors in the context of these analyses seemed to be small.

**Table 1.** Proportion of variance in R^2^ that can be explained by different variables using type II sum of squares from ANOVA.

| Variable | Proportion of explained variance |
| --- | --- |
| Ancestry of summary statistics or test data | 0.5510 |
| Choice of LD reference panel | 0.1650 |
| Age of test individuals | 0.1590 |
| N individuals used to calculate summary statistics | 0.0195 |
| Residuals (unexplained variance) | 0.1050 |

### PRS_BMI_ association with comorbid traits

To determine whether the PRS_BMI_ was associated with clinical comorbidities, we performed a Phenome-Wide Association Study (described more in Methods). Here, the PRS_BMI_ was tested for association with diagnosis codes (Phecodes) to evaluate whether the polygenic background for BMI associates with these clinical diagnoses. The PRS_BMI_ was significantly associated with several of the most frequent Phecodes in eMERGE, particularly in European adults (Figure 3a). As expected, obesity had the strongest association with PRS_BMI_ in all ancestry groups (p_EUR_ < 4.9×10^−324^; p_AFR_ = 5.17×10^−8^); this is a positive control. In European ancestry individuals, the best performing PRS_BMI_ was also significantly positively associated with type 2 diabetes (p_EUR_ = 1.04×10^−102^), essential hypertension (p_EUR_ = 7.12×10^−56^), coronary atherosclerosis (p_EUR_ = 3.61×10^−26^), hyperlipidemia (p_EUR_ = 4.38×10^−16^), depression (p_EUR_ = 1.95×10^−13^), hypercholesteremia (p_EUR_ = 3.64×10^−15^), asthma (p_EUR_ = 3.13×10^−13^), and diverticulosis (p_EUR_ = 0.0017). These associations were less statistically significant in African ancestry individuals, potentially due to the lower sample size, and many associations were no longer significant after Bonferroni correction. Only type 2 diabetes (p_AFR_ = 1.2×10^−5^) and coronary atherosclerosis (p_AFR_ = 0.001) were significantly associated with the PRS_BMI_ in African ancestry adults. We also looked at the prevalence of each condition per PRS quintile for the most significantly associated conditions (Figure 3b). The case prevalence generally increased in higher PRS_BMI_ quintile groups for conditions significantly associated with the PRS_BMI_, a trend matching the results we obtained from the association analysis. We performed similar analyses in teens and children but identified no statistically significant associations (results not shown). The much smaller sample sizes of the Phecodes in these age groups may have also contributed to the lack of statistically significant results – most of these diagnoses are adult-onset conditions.

**Figure 3.**
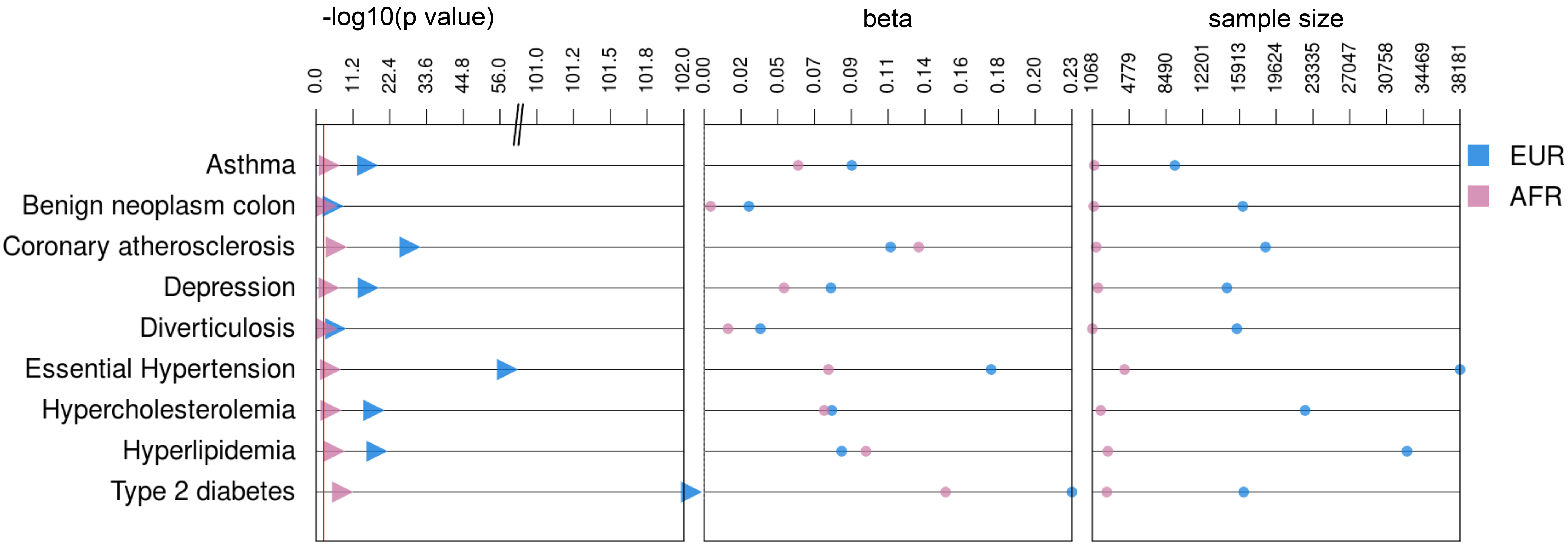
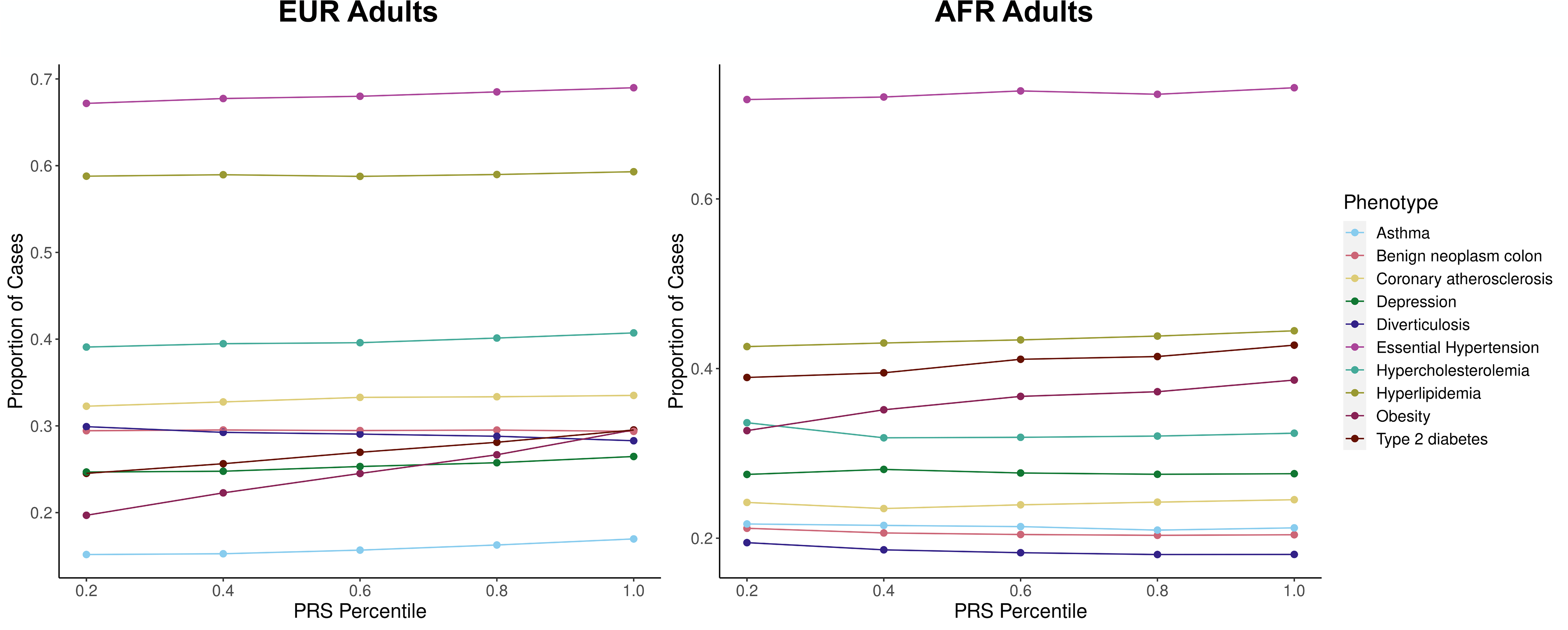
a) Best PRS_BMI_ associations with top 9 most prevalent conditions overall in eMERGE adults b) Prevalence plots of significantly associated conditions in eMERGE adults by best performing PRS quintile

## Discussion

Using the eMERGE network data, a diverse dataset in terms of ancestry and age, we were able to quantify the effects that different variables (specifically LD panel, age, ancestry, and sample size of GWAS summary statistics) have on PRS_BMI_ performance. We observed significant improvements in PRS_BMI_ performance for African ancestry individuals using a cross-ancestry GWAS to generate the PRS_BMI_ weights, compared to a European ancestry GWAS of the same size. Even for European ancestry individuals, we observed a significant increase in PRS_BMI_ performance when using a cross-ancestry GWAS versus a solely European ancestry GWAS despite both GWAS being the same sample size. These results suggest that cross-ancestry GWAS summary statistics significantly improve PRS_BMI_ performance not only for non-European ancestry individuals, but possibly for European ancestry individuals as well (or, at minimum, do not significantly degrade PRS_BMI_ performance for European ancestry individuals). Furthermore, when all variables were considered in a multivariate model, we observed that the ancestry of the GWAS summary statistics and test data affects the PRS_BMI_ performance the most. Despite the size of the GWAS having a significant effect on PRS_BMI_ performance, its overall impact is small compared to the other variables we considered in this study, suggesting that for PRS prediction in non-European ancestry individuals, the impact of increasing the sample size of European ancestry GWAS will be minimal compared to performing GWAS in individuals of diverse ancestry. This also suggests that it would be advantageous to use GWAS summary statistics from a similar ancestry group (or cross-ancestry GWAS) even if the sample size is modest, rather than a larger sample size European ancestry GWAS. These results are consistent with previous reports that LD and allele frequency differences explain most of the loss of relative accuracy in PRS performance between European and African ancestry individuals (23). These results also highlight the utility and importance of increasing the number of non-European ancestry individuals used in GWAS, as their inclusion disproportionately increases PRS performance in non-Europeans and may even increase PRS performance for European ancestry individuals. We also found that the PRS_BMI_ was positively associated with type 2 diabetes and coronary atherosclerosis in adults across ancestry groups, and we detected positive associations of the PRS_BMI_ with many other common comorbidities, such as essential hypertension and hyperlipidemia, for European ancestry adults.

Somewhat unintuitively, African ancestry LD panels performed best for African ancestry individuals, regardless of whether European ancestry or cross-ancestry GWAS summary statistics were used. We observed minimal impact of the choice of LD panel when both test data and summary statistics were of European ancestry. These results suggest that as long as either the test data or GWAS summary statistics are of similar ancestry, or the test data and LD panel are of similar ancestry, the difference in PRS performance may be minimal as compared to if all the GWAS summary statistics, test data, and LD panel are all of the same ancestry.

While the findings in this study highlight many important strategies for performing PRS in different ancestry groups, there are limitations that should be addressed in future studies. First, inclusion of analyses that evaluate how different proportions of non-European ancestry individuals affect the prediction performance of PRS would be useful. The GIANT summary statistics we used in this study are only about 6% non-European ancestry. It may be useful to see how the PRS prediction performance changes in both European and non-European datasets as a function of the proportion of non-European ancestry samples included in the GWAS. Such analyses may be possible by combining African ancestry individuals from these different datasets. In addition, similar analyses to evaluate, in a more robust manner, the impact of the overall size of the GWAS is greatly needed. These analyses will be possible once larger datasets that include non-European ancestry cohorts are publicly available.

Overall, this study demonstrates the importance of expanding non-European ancestry data resources for PRS, specifically in the generation of GWAS summary statistics and LD reference panels. Failure to do so reduces the impact of PRS in diverse populations and increase the potential for continued health disparities.

## Data Availability

All data produced in the present study are available upon reasonable request to the authors.

## Description of supplemental data

Supplemental data include one figure and two tables.

## Declaration of author competing interests

Authors have no competing interests to declare.

## Data and code availability

Code supporting the current study are available from the corresponding author on request.

## Notes

### Competing Interest Statement

The authors have declared no competing interest.

### Funding Statement

This work was in part supported by P50GM115318-04S1.
eMERGE Network (Phase III). This phase of the eMERGE Network was initiated and funded by the NHGRI through the following grants: U01HG8657 (Group Health Cooperative/University of Washington); U01HG8685 (Brigham and Womens Hospital); U01HG8672 (Vanderbilt University Medical Center); U01HG8666 (Cincinnati Childrens Hospital Medical Center); U01HG6379 (Mayo Clinic); U01HG8679 (Geisinger Clinic); U01HG8680 (Columbia University Health Sciences); U01HG8684 (Childrens Hospital of Philadelphia); U01HG8673 (Northwestern University); U01HG8701 (Vanderbilt University Medical Center serving as the Coordinating Center); U01HG8676 (Partners Healthcare/Broad Institute); and U01HG8664 (Baylor College of Medicine).
UK Biobank. All data for this cohort pertained to project 32133: Integration of multi-organ imaging phenotypes, clinical phenotypes, and genomic data.

### Author Declarations

This study was conducted under all relevant ethical regulations. UK Biobank was approved under application ID 32133. eMERGE study was approved under review of the board committee.

### Summary of Updates

Super and subscripts for abstract

## References

1. Truong B, Zhou X, Shin J, Li J, van der Werf JHJ, Le TD, et al. Efficient polygenic risk scores for biobank scale data by exploiting phenotypes from inferred relatives. Nat Commun. 2020 Jun 17;11(1):3074.

2. Margaux L.A. Hujoel, Po-Ru Loh, Benjamin M. Neale, Alkes L. Price. Incorporating family history of disease improves polygenic risk scores in diverse populations. Available from: https://www.biorxiv.org/content/10.1101/2021.04.15.439975v1

3. Martin AR, Kanai M, Kamatani Y, Okada Y, Neale BM, Daly MJ. Clinical use of current polygenic risk scores may exacerbate health disparities. Nat Genet. 2019 Apr;51(4):584–91.

4. Rask-Andersen M, Karlsson T, Ek WE, Johansson Å. Gene-environment interaction study for BMI reveals interactions between genetic factors and physical activity, alcohol consumption and socioeconomic status. PLoS Genet. 2017 Sep;13(9):e1006977.

5. Robinson MR, English G, Moser G, Lloyd-Jones LR, Triplett MA, Zhu Z, et al. Genotype-covariate interaction effects and the heritability of adult body mass index. Nat Genet. 2017 Aug;49(8):1174–81.

6. Sulc J, Mounier N, Günther F, Winkler T, Wood AR, Frayling TM, et al. Quantification of the overall contribution of gene-environment interaction for obesity-related traits. Nat Commun. 2020 Mar 13;11(1):1385.

7. Justice AE, Winkler TW, Feitosa MF, Graff M, Fisher VA, Young K, et al. Genome-wide meta-analysis of 241,258 adults accounting for smoking behaviour identifies novel loci for obesity traits. Nat Commun. 2017 Apr 26;8:14977.

8. Helgeland Ø, Vaudel M, Juliusson PB, Lingaas Holmen O, Juodakis J, Bacelis J, et al. Genome-wide association study reveals dynamic role of genetic variation in infant and early childhood growth. Nat Commun. 2019 Oct 1;10(1):4448.

9. Vogelezang S, Bradfield JP, Ahluwalia TS, Curtin JA, Lakka TA, Grarup N, et al. Novel loci for childhood body mass index and shared heritability with adult cardiometabolic traits. PLoS Genet. 2020 Oct;16(10):e1008718.

10. Couto Alves A, De Silva NMG, Karhunen V, Sovio U, Das S, Taal HR, et al. GWAS on longitudinal growth traits reveals different genetic factors influencing infant, child, and adult BMI. Sci Adv. 2019 Sep;5(9):eaaw3095.

11. Choh AC, Lee M, Kent JW, Diego VP, Johnson W, Curran JE, et al. Gene-by-age effects on BMI from birth to adulthood: the Fels Longitudinal Study. Obes Silver Spring Md. 2014 Mar;22(3):875–81.

12. Mostafavi H, Harpak A, Agarwal I, Conley D, Pritchard JK, Przeworski M. Variable prediction accuracy of polygenic scores within an ancestry group. eLife. 2020 Jan 30;9:e48376.

13. Elks CE, den Hoed M, Zhao JH, Sharp SJ, Wareham NJ, Loos RJF, et al. Variability in the heritability of body mass index: a systematic review and meta-regression. Front Endocrinol. 2012;3:29.

14. Galinsky KJ, Reshef YA, Finucane HK, Loh P-R, Zaitlen N, Patterson NJ, et al. Estimating cross-population genetic correlations of causal effect sizes. Genet Epidemiol. 2019 Mar;43(2):180–8.

15. Shi H, Gazal S, Kanai M, Koch EM, Schoech AP, Siewert KM, et al. Population-specific causal disease effect sizes in functionally important regions impacted by selection. Nat Commun. 2021 Feb 17;12(1):1098.

16. Stanaway IB, Hall TO, Rosenthal EA, Palmer M, Naranbhai V, Knevel R, et al. The eMERGE genotype set of 83,717 subjects imputed to ∼40 million variants genome wide and association with the herpes zoster medical record phenotype. Genet Epidemiol. 2019 Feb;43(1):63–81.

17. Locke AE, Kahali B, Berndt SI, Justice AE, Pers TH, Day FR, et al. Genetic studies of body mass index yield new insights for obesity biology. Nature. 2015 Feb 12;518(7538):197–206.

18. Bycroft C, Freeman C, Petkova D, Band G, Elliott LT, Sharp K, et al. The UK Biobank resource with deep phenotyping and genomic data. Nature. 2018 Oct;562(7726):203–9.

19. Howie BN, Donnelly P, Marchini J. A flexible and accurate genotype imputation method for the next generation of genome-wide association studies. PLoS Genet. 2009 Jun;5(6):e1000529.

20. Euesden J, Lewis CM, O’Reilly PF. PRSice: Polygenic Risk Score software. Bioinforma Oxf Engl. 2015 May 1;31(9):1466–8.

21. Wu P, Gifford A, Meng X, Li X, Campbell H, Varley T, et al. Mapping ICD-10 and ICD-10-CM Codes to Phecodes: Workflow Development and Initial Evaluation. JMIR Med Inform. 2019 Nov 29;7(4):e14325.

22. Pendergrass SA, Dudek SM, Crawford DC, Ritchie MD. Visually integrating and exploring high throughput Phenome-Wide Association Study (PheWAS) results using PheWAS-View. BioData Min. 2012 Jun 8;5(1):5.

23. Wang Y, Guo J, Ni G, Yang J, Visscher PM, Yengo L. Theoretical and empirical quantification of the accuracy of polygenic scores in ancestry divergent populations. Nat Commun. 2020 Jul 31;11(1):3865.

