## Supplemental for "Quantifying factors that affect polygenic risk score performance across diverse ancestries and age groups for body mass index"

S Table 1. Effect of GWAS sample size on PRS performance. Difference in R^2^ for different groupings of individuals between the UKBB GWAS down-sampled to that of the sample size used in GIANT (N=226,960), and the full UKBB European sample size (N=377,921). Test African ancestry individuals and 5,000 random European ancestry UKBB individuals were used as LD panels for all African and European ancestry test sets, respectively.

| Age group | Ancestry | N | PRS R^2^ UKBB  Down-sampled sample size | PRS R^2^  UKBB  Full sample size | P-value |
| --- | --- | --- | --- | --- | --- |
| Adults | European | 55418 | 0.0582 | 0.0734 | <4.94x10^-324^ |
| Teenagers |  | 3114 | 0.0588 | 0.0667 | 3.75x10^-9^ |
| Children |  | 5943 | 0.0274 | 0.0347 | 6.51x10^-47^ |
| Adults | African | 5912 | 0.0140 | 0.0197 | 1.85x10^-46^ |
| Teenagers |  | 1606 | 0.0179 | 0.0274 | 1.37x10^-29^ |
| Children |  | 3668 | 0.00612 | 0.00715 | 4.55x10^-9^ |

S Table 2. Selected Phecodes in eMERGE which were tested for their association to the PRS_BMI_.

| Phecode | Description | Cases/Controls (Adults) | |
| --- | --- | --- | --- |
|  |  | EUR | AFR |
| 401.1 | Essential hypertension | 38181/17166 | 4317/1588 |
| 272.1 | Hyperlipidemia | 32823/22524 | 2625/3280 |
| 272.11 | Hypercholesterolemia | 22537/32810 | 1912/3993 |
| 278.1 | Obesity (positive control) | 16348/38999 | 2282/3623 |
| 411.4 | Coronary atherosclerosis | 18548/36799 | 1449/4456 |
| 250.2 | Type 2 diabetes | 16342/39005 | 2525/3380 |
| 296.2 | Depression | 14651/40696 | 1630/4275 |
| 208 | Benign neoplasm of colon | 16254/39093 | 1205/4700 |
| 562.1 | Diverticulosis | 15656/39691 | 1068/4837 |
| 495 | Asthma | 9391/45956 | 1253/4652 |

S Figure 1. Differences in PRS R^2^ across different age groups (the best performing LD panel was used for each run, p-values above each bar). We consistently observed more significant differences in PRS R^2^ in children versus adults and teenagers. The down-sampled UKBB summary statistics were used for these comparisons.

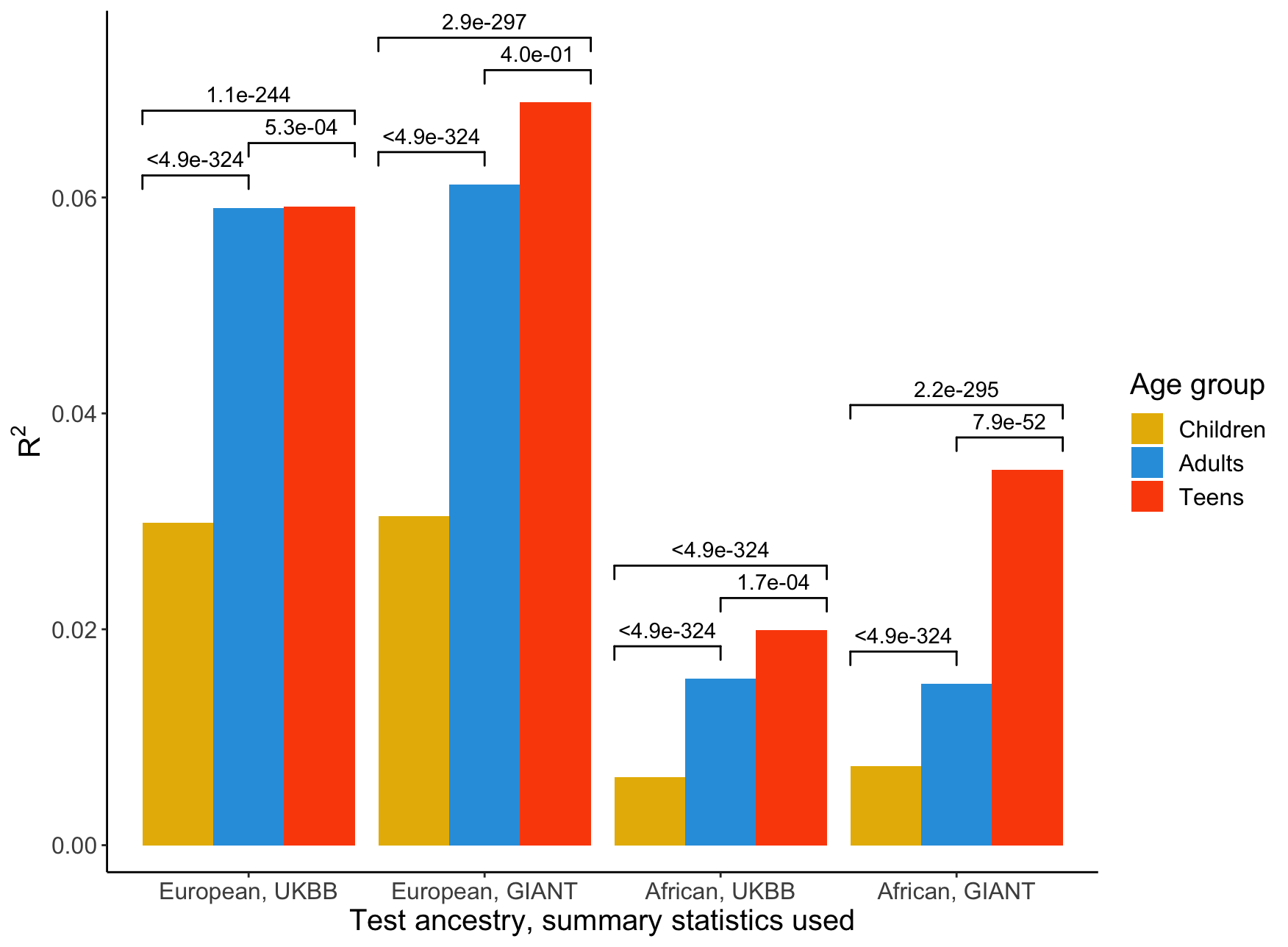
